# Clinical Subphenotypes of Multisystem Inflammatory Syndrome in Children: An EHR-based cohort study from the RECOVER program

**DOI:** 10.1101/2022.09.26.22280364

**Authors:** Suchitra Rao, Naimin Jing, Xiaokang Liu, Vitaly Lorman, Mitchell Maltenfort, Julia Schuchard, Qiong Wu, Jiayi Tong, Hanieh Razzaghi, Asuncion Mejias, Grace M. Lee, Nathan M. Pajor, Grant S. Schulert, Deepika Thacker, Ravi Jhaveri, Dimitri A. Christakis, L. Charles Bailey, Christopher B. Forrest, Yong Chen, the RECOVER consortium

## Abstract

**Background:** Multi-system inflammatory syndrome in children (MIS-C) represents one of the most severe post-acute sequelae of SARS-CoV-2 infection in children, and there is a critical need to characterize its disease patterns for improved recognition and management. Our objective was to characterize subphenotypes of MIS-C based on presentation, demographics and laboratory parameters.

**Methods:** We conducted a retrospective cohort study of children with MIS-C from March 1, 2020 - April 30, 2022 and cared for in 8 pediatric medical centers that participate in PEDSnet. We included demographics, symptoms, conditions, laboratory values, medications and outcomes (ICU admission, death), and grouped variables into eight categories according to organ system involvement. We used a heterogeneity-adaptive latent class analysis model to identify three clinically-relevant subphenotypes. We further characterized the sociodemographic and clinical characteristics of each subphenotype, and evaluated their temporal patterns.

**Findings:** We identified 1186 children hospitalized with MIS-C. The highest proportion of children (44·4%) were aged between 5-11 years, with a male predominance (61.0%), and non- Hispanic white ethnicity (40·2%). Most (67·8%) children did not have a chronic condition. Class 1 represented children with a severe clinical phenotype, with 72·5% admitted to the ICU, higher inflammatory markers, hypotension/shock/dehydration, cardiac involvement, acute kidney injury and respiratory involvement. Class 2 represented a moderate presentation, with 4-6 organ systems involved, and some overlapping features with acute COVID-19. Class 3 represented a mild presentation, with fewer organ systems involved, lower CRP, troponin values and less cardiac involvement. Class 1 initially represented 51·1% of children early in the pandemic, which decreased to 33·9% from the pre-delta period to the omicron period.

**Interpretation:** MIS-C has a spectrum of clinical severity, with degree of laboratory abnormalities rather than the number of organ systems involved providing more useful indicators of severity. The proportion of severe/critical MIS-C decreased over time.

**Research in context:** *Evidence before this study:* We searched PubMed and preprint articles from December 2019, to July 2022, for studies published in English that investigated the clinical subphenotypes of MIS-C using the terms “multi-system inflammatory syndrome in children” or “pediatric inflammatory multisystem syndrome” and “phenotypes”. Most previous research described the symptoms, clinical characteristics and risk factors associated with MIS-C and how these differ from acute COVID-19, Kawasaki Disease and Toxic Shock Syndrome. One single-center study of 63 patients conducted in 2020 divided patients into Kawasaki and non-Kawasaki disease subphenotypes. Another CDC study evaluated 3 subclasses of MIS-C in 570 children, with one class representing the highest number of organ systems, a second class with predominant respiratory system involvement, and a third class with features overlapping with Kawasaki Disease. However, this study evaluated cases from March to July 2020, during the early phase of the pandemic when misclassification of cases as Kawasaki disease or acute COVID-19 may have occurred. Therefore, it is not known from the existing literature whether the presentation of MIS-C has changed with newer variants such as delta and omicron.

*Added value of this study:* PEDSnet provides one of the largest MIS-C cohorts described so far, providing sufficient power for detailed analyses on MIS-C subphenotypes. Our analyses span the entire length of the pandemic, including the more recent omicron wave, and provide an update on the presentations of MIS-C and its temporal dynamics. We found that children have a spectrum of illness that can be characterized as mild (lower inflammatory markers, fewer organ systems involved), moderate (4-6 organ involvement with clinical overlap with acute COVID-19) and severe (higher inflammatory markers, critically ill, more likely to have cardiac involvement, with hypotension/shock and need for vasopressors).

*Implications of all the available evidence:* These results provide an update to the subphenotypes of MIS-C including the more recent delta and omicron periods and aid in the understanding of the various presentations of MIS-C. These and other findings provide a useful framework for clinicians in the recognition of MIS-C, identify factors associated with children at risk for increased severity, including the importance of laboratory parameters, for risk stratification, and to facilitate early evaluation, diagnosis and treatment.

## Introduction

In April 2020, cases of a novel hyperinflammatory disorder associated with SARS-CoV-2 infection were first reported in the UK and Italy,^1,2^ which was subsequently termed multisystem inflammatory syndrome in children (MIS-C). The Centers for Disease Control and Prevention (CDC) defines cases as individuals younger than 21 years of age presenting with fever, laboratory evidence of inflammation, and evidence of clinically severe illness requiring hospitalization, with involvement of greater than or equal to 2 organs, with no alternative plausible diagnoses and a positive SARS-CoV-2 test (PCR, serology, antigen) or COVID-19 exposure within the preceding 4 weeks.^3^ Reports indicate that most children are 6-12 years of age, of Black/African American race or Hispanic ethnicity, and obesity was the most common pre-existing condition.^4-6^ MIS-C is one of the most severe post-acute sequelae of SARS-CoV-2 infection in children, and as of June 2022, the CDC has reported 8,639 cases of MIS-C; 70 children have died due to associated complications.^7^

Children with MIS-C can present with a highly heterogeneous spectrum of clinical features, making the diagnosis challenging. The clinical presentation can overlap with features of Kawasaki disease, acute COVID-19, toxic shock, and macrophage activation syndrome. Common features include fever, gastrointestinal symptoms (including abdominal pain and diarrhea), cardiac complications (myocarditis and coronary artery dilatation), mucocutaneous findings (rash, mucositis) and respiratory symptoms.^8^ Given the wide spectrum of clinical presentations of this novel condition, there is a need to further characterize its disease patterns, which will ultimately improve outcomes. Further elucidation of MIS-C subphenotypes can help identify children at greatest risk of severe outcomes, and determine which subgroups may benefit from more targeted therapies. Therefore, the objectives of this study were to define subphenotypes of MIS-C based on symptoms, diagnoses and laboratory parameters.

## Methods

### Data Source

This retrospective cohort study is part of the NIH Researching COVID to Enhance Recovery (RECOVER) Initiative, which seeks to understand, treat, and prevent the post-acute sequelae of SARS-CoV-2 infection (PASC). For more information on RECOVER, visit https://recovercovid.org/. The original study cohort has been described elsewhere.^9^ We used electronic health record (EHR) data from PEDSnet member institutions.^10,11^ Participating institutions included Children’s Hospital of Philadelphia, Cincinnati Children’s Hospital Medical Center, Children’s Hospital Colorado, Ann & Robert H. Lurie Children’s Hospital of Chicago, Nationwide Children’s Hospital, Nemours Children’s Health System (a Delaware and Florida health system), Seattle Children’s Hospital, and Stanford Children’s Health. We retrieved EHR data from all healthcare encounters among hospitalized children and adolescents < 21 years of age who were diagnosed with MIS-C from March 1, 2020 to April 30, 2022. Data were extracted from the PEDSnet COVID-19 Database-Version 2022-05-19, which included EHR data with dates of service up to April 30, 2022. Cohort entry was defined as the 1 day before the first hospitalization associated with an MIS-C diagnosis term (index hospitalization). We combined hospitalizations if the discharge date of the first hospitalization was within 24 hours of the subsequent hospitalization admission date. We excluded one patient with missing admission or discharge dates. The Children’s Hospital of Philadelphia’s Institutional Review Board designated this study as not human subjects’ research and waived informed consent.

### Variables

We included the following indicator variables: demographics, diagnoses (including symptoms and conditions), medications, laboratory values and clinical outcomes (ICU, mechanical ventilation, death), which were coded dichotomously and later grouped to form variables used in our analysis. The indicator variable was included in the analyses if at least one occurrence was recorded in the EHR from 1 day prior through the entire duration of hospitalization. In addition, age, number of days during hospitalization, number of days in ICU, and laboratory test results were included as continuous or categorical variables.

### Statistical Analyses

First, we grouped highly-correlated indicator variables, including diagnoses (symptoms and conditions) and laboratory values, into eight broader categories according to their organ system involvement, which were cardiac, gastrointestinal, hematological, neurologic, renal, respiratory, dermatologic and shock (See **eTable 1** for definitions of each variable). The variable for a particular organ system was defined based on the occurrence of any symptom, condition, laboratory value, or procedure type related to that system. We further divided the cardiac system into three categories: coronary artery abnormalities only, myocardial dysfunction with no coronary artery abnormalities, and coronary artery abnormalities and myocardial dysfunction.

Secondly, with the ten manifest variables defined over the eight broad organ systems, we applied a latent class analysis (LCA) model in which an Expectation-Maximization (EM) algorithm was used for estimation.^12,13^ To account for heterogeneity across hospitals yet ensuring unified definitions of subphenotypes, we assumed the same collection of subphenotypes but potentially different mixing proportions of each of the subphenotypes across the health systems. Such generalization of the traditional LCA model is necessary because the prevalence of subphenotypes can be substantially different due to different distributions of age, race/ethnicity, and referral patterns which can be strongly associated with the presentation of MIS-C.

For sensitivity analysis, we fitted LCA to the ten manifest variables under different assumed numbers of latent classes, varying from two to six. Log-likelihood, Bayesian Information Criterion (BIC), and clinical adjudication among our clinician work group were incorporated to select the appropriate number of clinical-relevant latent classes.

For identification of identified latent classes (subphenotypes), proportions of the ten manifest variables over the eight broad organ systems across latent classes were displayed as heatmaps of prevalence. To further characterize these identified latent classes, distributions of clinical indicator variables, including demographic characteristics, and individual symptoms, conditions, laboratory results, procedures, and medications, were displayed across the latent classes also using heatmaps of prevalence. To characterize the differences in each continuous clinical variable across latent classes, sample mean, standard error, median and interquartile range were calculated and summarized. To reduce the impact of misclassifications of latent classes, instead of using classification based on maximum posterior probability, all calculations of the characterizations were based on weighted prevalence with weights being the posterior probabilities. The results are presented using heatmaps and forest plots.

To evaluate potential time-varying effects, we also evaluated the changes of the proportions for each subphenotype over time in 3-month periods. Within each time period, the proportions of subphenotypes were calculated using posterior class membership probabilities. The change in subclass proportions over time was shown as a line graph in Figure 4. In addition, the change in the prevalence of the ten variables for each of the latent classes over time was estimated.

To further examine the validity of our LCA approach, we applied the subphenotypes of MIS-C to a cohort of children testing positive for COVID-19 by PCR without an MIS-C diagnosis.^9^ We constructed latent classes for SARS-CoV-2 positive patients, with an evaluation period of −7 to 28 days from the test date to evaluate our construct validity. We applied the model with three latent classes to the ten variables as used in the MIS-C analysis and identified three subphenotypes of COVID-19. We compared the characteristics of MIS-C and COVID-19 subphenotypes and visualized the distance among them measured by fixation index using multidimensional scaling. Analyses were conducted using R version 4.1.2 (2021-11-01). The package used for LCA was poLCA 1.4.1 (2014-01-10).^14,15^

## Results

Among 1,853,557 children in the PEDSnet cohort who received a COVID-19 diagnosis or vaccination, had a COVID-19 test (PCR, serology or antigen), or demonstrated evidence of respiratory illness or post-acute sequelae of SARS-CoV-2 and had a visit at any of the eight PEDSnet sites between March 2020 and April 2022, we identified 1186 children who were hospitalized with MIS-C (**eFigure 1**). A description of the study cohort is provided in **Table 1**. The highest proportion of children (44%) were aged between 5-11 years, with a male predominance (61%), and non-Hispanic White ethnicity (40·2%). Most (67·8%) children did not have any chronic conditions as defined by the Pediatric Medical Complexity Algorithm (PMCA). ^16^ In our MIS-C cohort, 40.5% were admitted to the ICU, 15.2% required mechanical ventilation (invasive and non-invasive), and 11 (0·92%) children died.

**Table 1.**
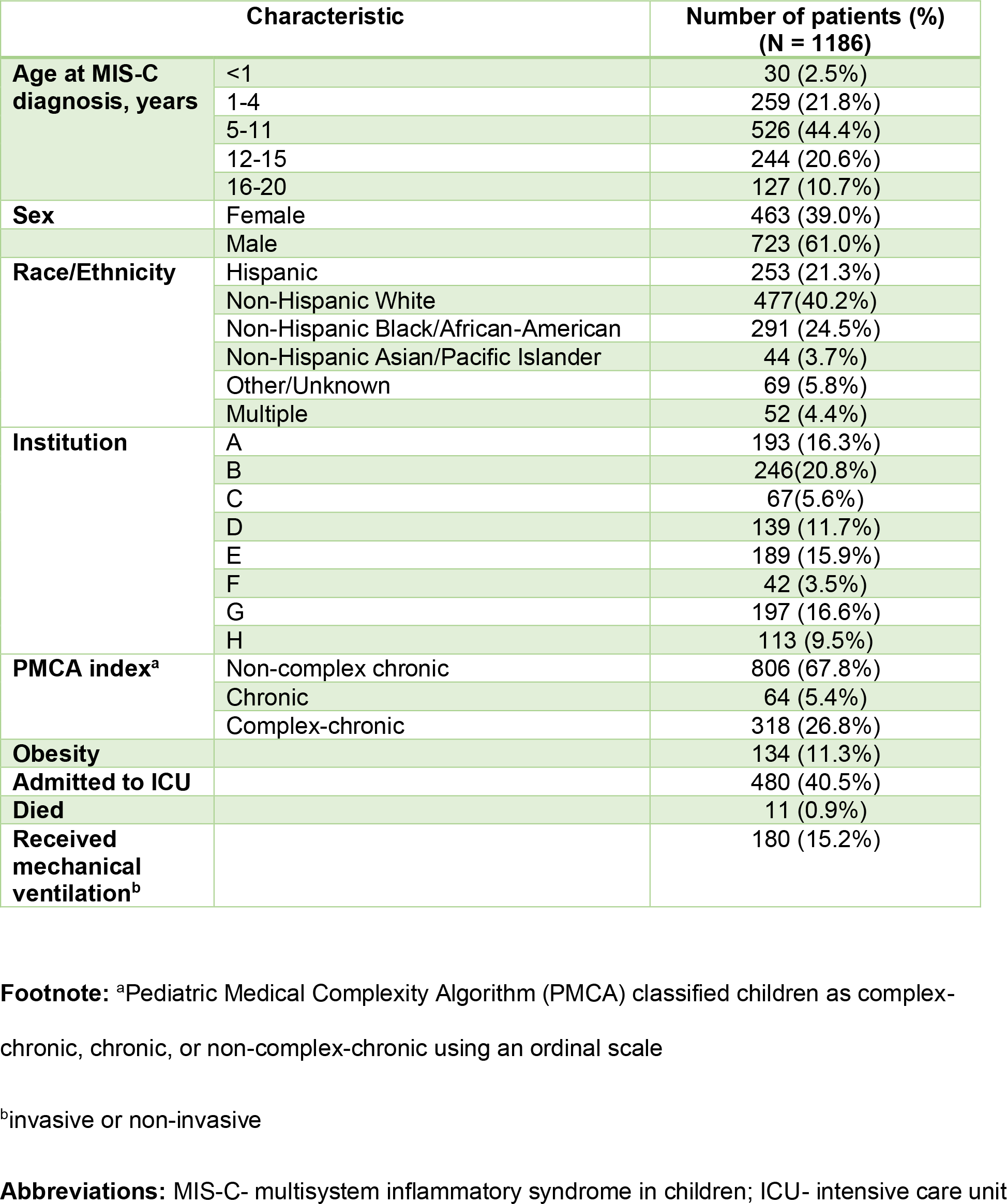
Description of Study Cohort.

| Characteristic |  | Number of patients (%)<br>(N = 1186) |
| --- | --- | --- |
| Age at MIS-C diagnosis, years | <1 | 30 (2.5%) |
|  | 1-4 | 259 (21.8%) |
|  | 5-11 | 526 (44.4%) |
|  | 12-15 | 244 (20.6%) |
|  | 16-20 | 127 (10.7%) |
| Sex | Female | 463 (39.0%) |
|  | Male | 723 (61.0%) |
| Race/Ethnicity | Hispanic | 253 (21.3%) |
|  | Non-Hispanic White | 477 (40.2%) |
|  | Non-Hispanic Black/African-American | 291 (24.5%) |
|  | Non-Hispanic Asian/Pacific Islander | 44 (3.7%) |
|  | Other/Unknown | 69 (5.8%) |
|  | Multiple | 52 (4.4%) |
| Institution | A | 193 (16.3%) |
|  | B | 246 (20.8%) |
|  | C | 67 (5.6%) |
|  | D | 139 (11.7%) |
|  | E | 189 (15.9%) |
|  | F | 42 (3.5%) |
|  | G | 197 (16.6%) |
|  | H | 113 (9.5%) |
| PMCA index<sup>a</sup> | Non-complex chronic | 806 (67.8%) |
|  | Chronic | 64 (5.4%) |
|  | Complex-chronic | 318 (26.8%) |
| Obesity |  | 134 (11.3%) |
| Admitted to ICU |  | 480 (40.5%) |
| Died |  | 11 (0.9%) |
| Received mechanical ventilation<sup>b</sup> |  | 180 (15.2%) |

**Footnote:** <sup>a</sup>Pediatric Medical Complexity Algorithm (PMCA) classified children as complex-chronic, chronic, or non-complex-chronic using an ordinal scale
<sup>b</sup>invasive or non-invasive
**Abbreviations:** MIS-C- multisystem inflammatory syndrome in children; ICU- intensive care unit

Latent Class analyses identified three subphenotypes (**Figure 1**), with the patient demographics of each class summarized in **Figure 2 and eFigure 2**. The overall proportions of Class 1, 2, and 3 were 40·0%, 24·0%, and 36·0%, respectively. The race and ethnicity of children were similar in each class, with the exception of a higher proportion of children of black/African American race in Class 1. A higher proportion of children 12 years of age and older, and those with a complex chronic condition were identified in Class 1. The proportion of each class by PEDSnet site is shown in **eTable 2**, with some observed heterogeneity. **Figure 3 and eFigure 3** and display specific diagnoses, laboratory results and medications in each group. Class 1 represented children with a more severe clinical phenotype, with 72·5% admitted to the ICU, with a longer mean inpatient stay and longer ICU length of stay. Children in Class 1 had a higher prevalence of cardiac involvement, with abnormal troponin levels, hypotension and shock/dehydration, need for vasopressors and fluid resuscitation. This class was more likely to have abnormal laboratory findings including lymphopenia, elevated d-dimer and thrombocytopenia. There was a higher prevalence of acute kidney injury, and respiratory involvement (respiratory failure, pleural effusion, need for ventilation). Children in Class 1 had the highest usage of systemic steroids among all classes. Class 2 represented a group with a moderate presentation, with similar multiple organ system involvement to Class 3, including dermatologic (rashes), gastrointestinal (abdominal pain, nausea and vomiting), hematologic abnormalities (lymphopenia, thrombocytopenia, elevated d-dimer), renal (fluid and electrolyte) and respiratory (cardiorespiratory signs and symptoms), with a heat map pattern similar to acute COVID-19 (**eFigure 4)** and 2-dimensional plot demonstrating close proximity between MIS-C Class 2 and COVID-19 Class 1 **(eFigure 5**). This group had the highest use of non-steroidal anti-inflammatory and antirheumatic medications. Class 3 represented a mild population, with fewer systems involved, and laboratory values demonstrating fluid and electrolyte disturbance, lymphopenia, thrombocytopenia, and elevated d-dimer. The proportions of each subphenotype over time are shown in **Figure 4**. A higher proportion of children had the more severe MIS-C presentation in the earlier phase of the pandemic (Class 1 represented 51·1% of children from March 2020 to August 2020), which decreased over time (Class 1 represented 33·9% of children from January to April, 2022). **eFigure 6** shows the prevalence of organ system involvement for all patients with MIS-C over time, demonstrating significant declines over time in cardiac, hematological, respiratory involvement, and shock.

**Figure 1.**
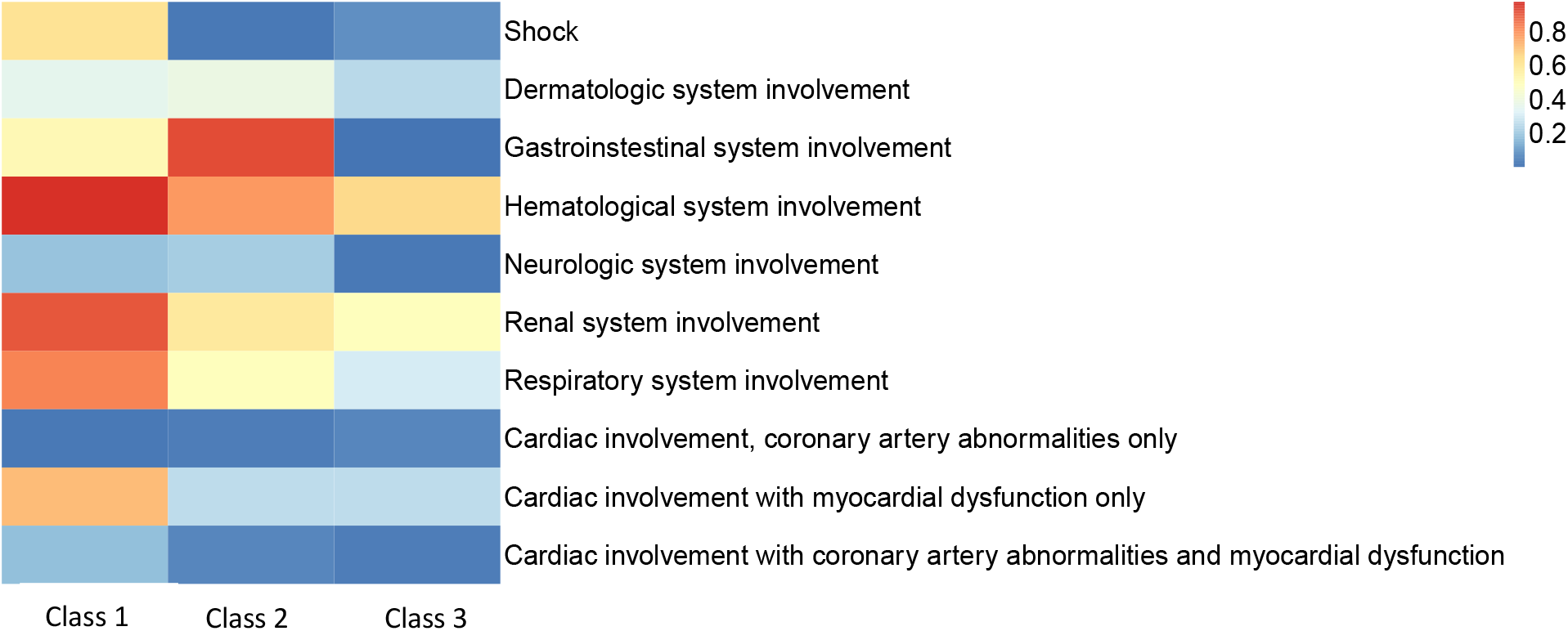
Heatmap of eight organ system variables used in latent class analyses with three classes. The proportions of Class 1, 2, and 3 are 40·0%, 24·0%, and 36·0%, respectively. Each column represents a latent class, and each row represents a manifest variable. The color of the boxes represents the prevalence of variables. The legend on the top right shows the scale of the colors. Red represents prevalence close to 100% and blue represents prevalence close to 0%.

**Figure 2.**
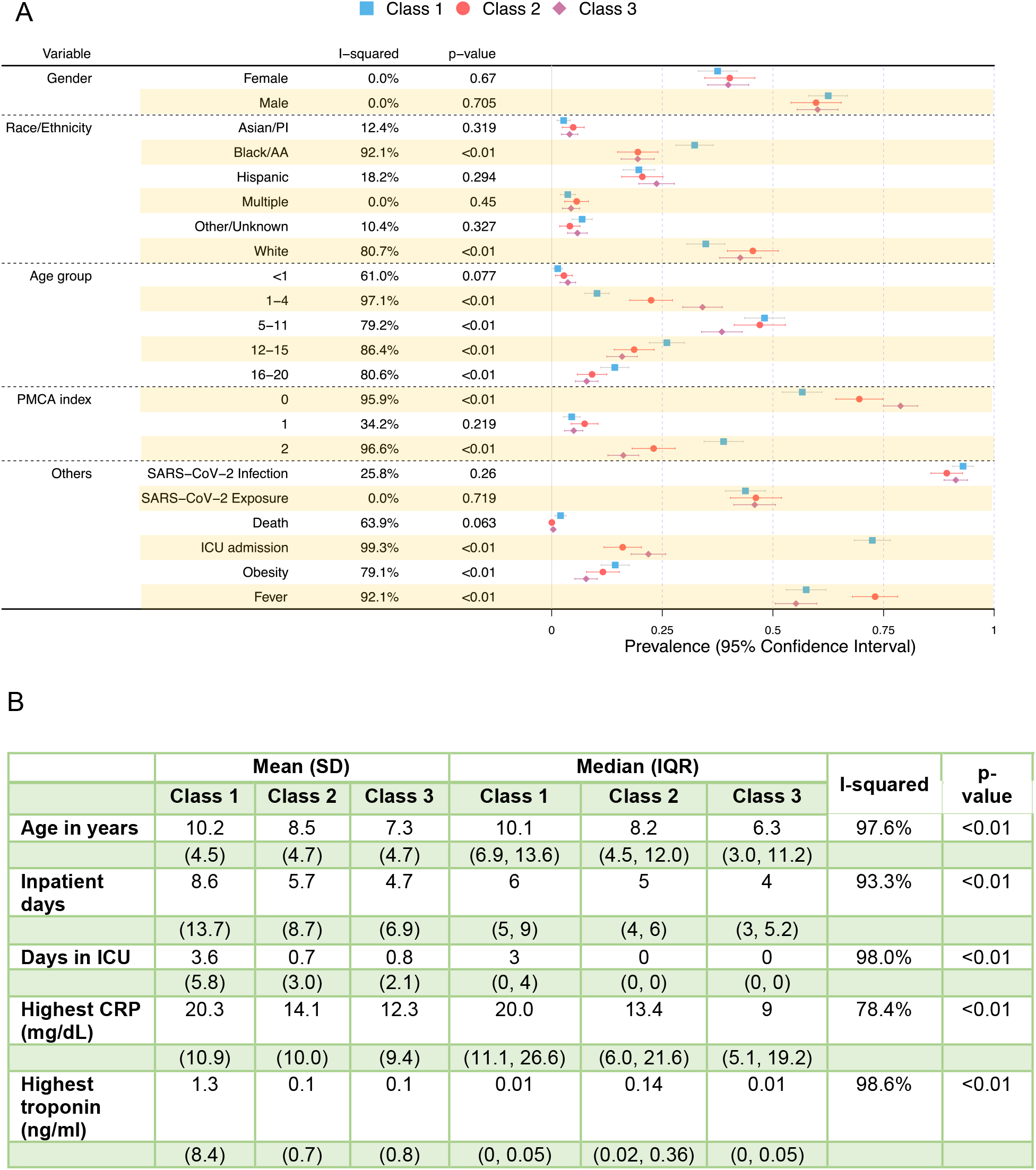
Patient demographic characteristics in three latent classes. Each line plot indicates the estimated prevalence and associated 95% confidence interval of each variable in each class. Class 1 is represented as a blue square, Class 2 as a red circle, and Class 3 as a purple diamond. The I-squared statistic measures the heterogeneity among the latent classes. P-values are obtained through Cochran’s Q test, where the null hypothesis is no between-class heterogeneity. A large I-squared and a small p-squared indicate a large between-class heterogeneity. **A)** I-squared, p-value, estimated prevalence, and its 95% confidence interval for discrete variables. **B)** I-squared, p-value, estimated mean, standard deviation, median and interquartile range for continuous variables. **Footnote:** SARS-CoV-2 infection includes positive PCR, antigen or serology; SARS-CoV-2 exposure indicates that patient had a history of exposure to a contact with known SARS-CoV-2 based on ICD-10 codes; age group is presented in years; Pediatric Medical Complexity Algorithm (PMCA) classified children as complex-chronic (2), chronic (1), or non-complex-chronic (0). **Abbreviations:** SD--standard deviation; IQR—interquartile range; PMCA-- Pediatric Medical Complexity Algorithm; PI- Pacific Islander; AA- African American; SARS-CoV-2--severe acute respiratory syndrome coronavirus; ICU- intensive care unit; CRP- C-reactive protein

**Figure 3.**
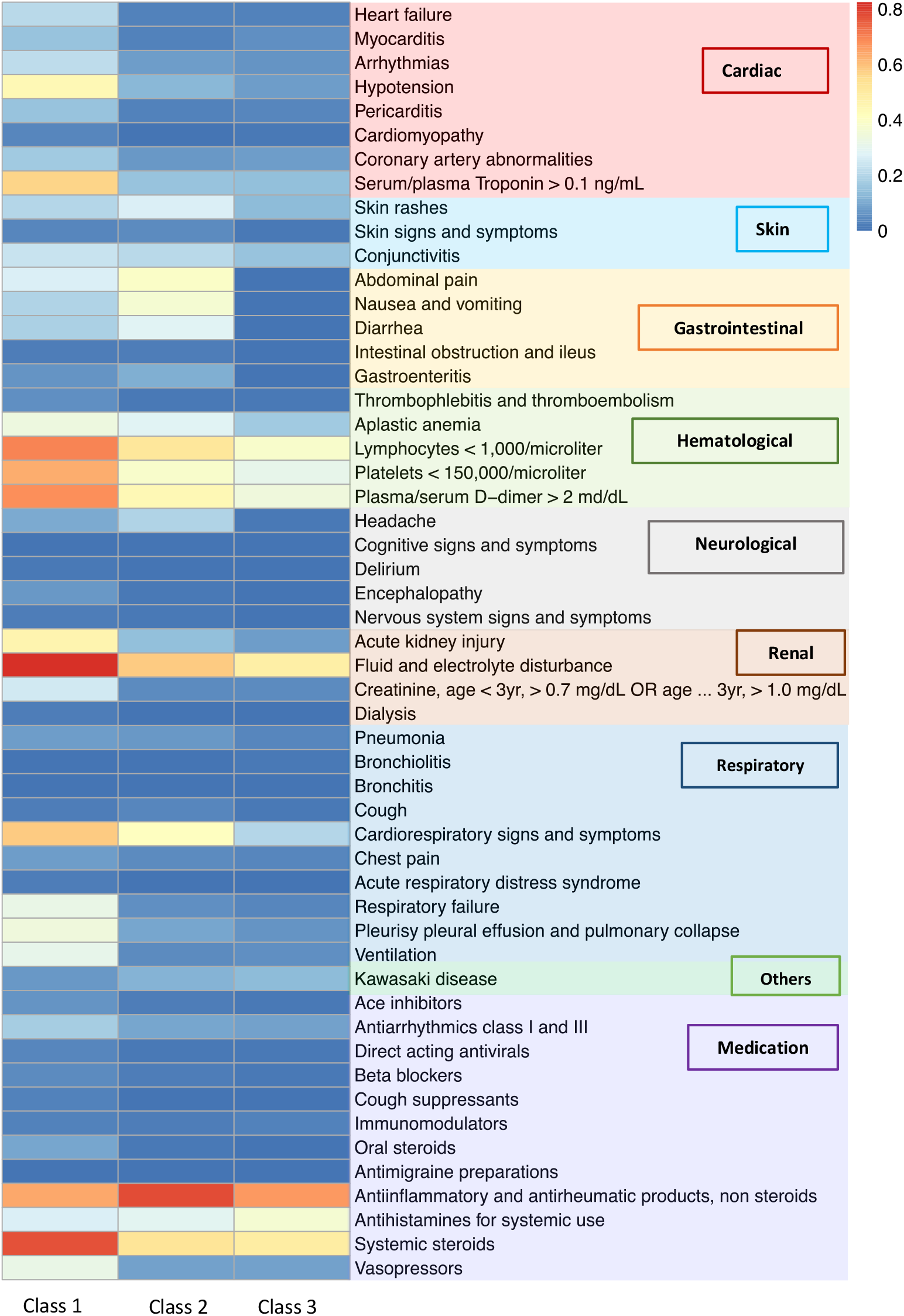
Heatmap of patient characteristics of conditions, laboratory results and medication in three latent classes. Each column represents a latent class, and each row represents a variable. The color of the boxes represents the estimated prevalence of variables. The legend on the top right shows the scale of the colors. Red represents prevalence close to 100% and blue represents prevalence close to 0%. The conditions and laboratory results are ordered based on organ systems.

**Figure 4.**
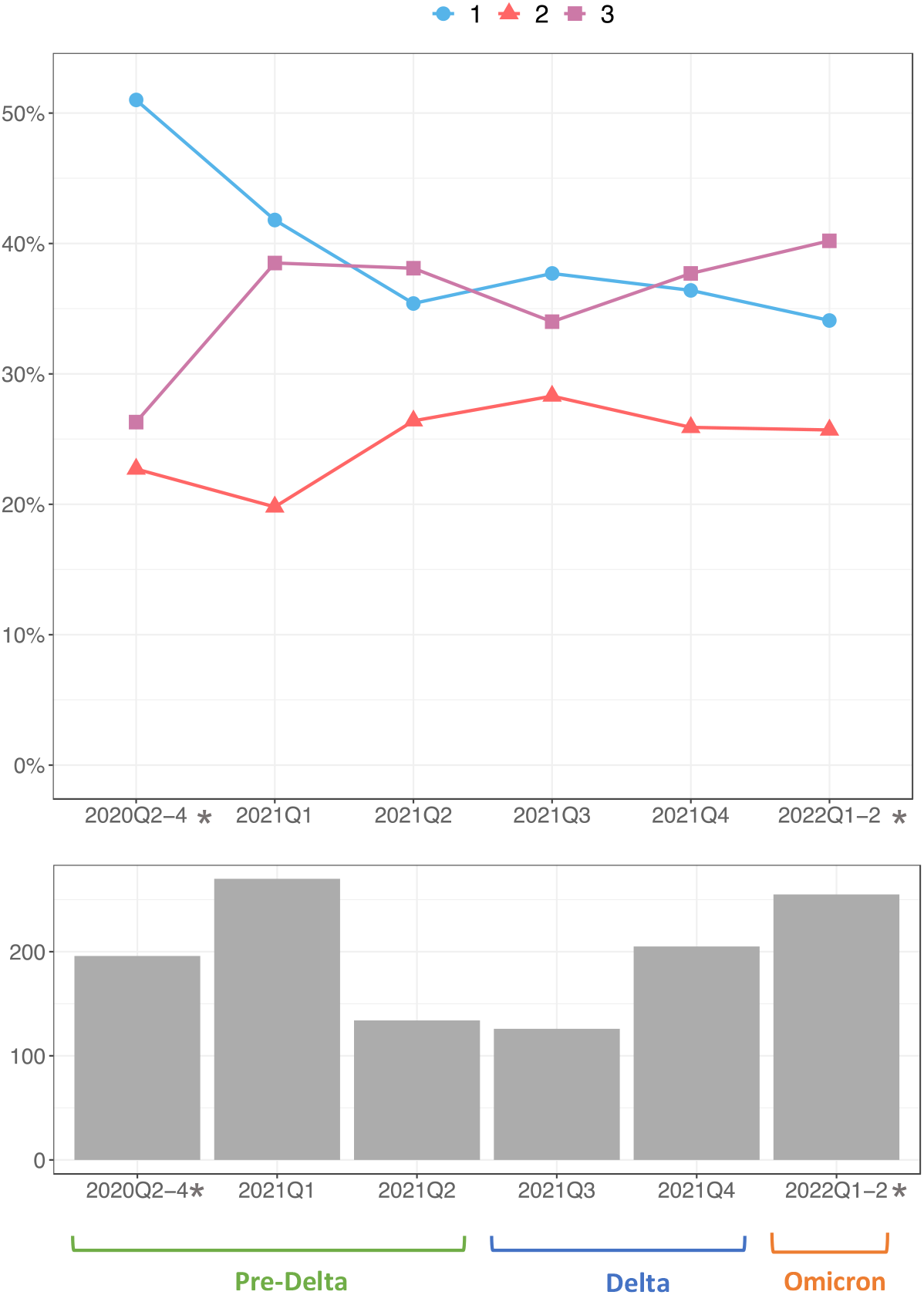
Proportions of three MIS-C classes over time. Each line represents the proportion of the three classes over 3-month periods. The proportions of the classes are calculated using the average of the posterior class membership probabilities of the patients who are diagnosed in the given time window. The calendar time is segmented quarterly. Time intervals with small sample sizes are combined and labeled by a star. **A)** Proportions of the three subphenotypes across time. Each line represents a subphenotype. **B)** Histogram of the number of patients in each time window, including timing of predominant SARS-CoV-2 variant activity.

## Discussion

In our multicenter cohort of 1,186 children hospitalized with MIS-C, we identified 3 latent classes with different illness characteristics. Class 1 represented critically ill patients, with a higher proportion of children being of older age, black/African American race, with underlying medical complexity, presenting with hypotension/shock, dehydration, cardiac dysfunction and laboratory abnormalities including lymphopenia, thrombocytopenia, elevated d-dimer and troponin. Class 3 represented a younger population with the milder form of MIS-C, and fewer organ systems involved (hematological and renal). Class 2 represented an intermediate population with lower illness severity than Class 1 but greater than Class 3, with 4-5 organ system involvement, including gastrointestinal (nausea, vomiting, abdominal pain, diarrhea), respiratory, hematological and renal, and a lower incidence of cardiac dysfunction and abnormal troponin levels, with a similar pattern to a subset of children with acute COVID-19. We also found a higher proportion of patients with the more severe Class 1 phenotype seen earlier in the pandemic, which decreased over time, with lower proportions of children with respiratory, cardiac and hematological involvement and shock. These findings highlight the varied presentations of children with MIS-C, with wide illness spectrum, variable system involvement, and cardiac involvement mostly confined to myocardial dysfunction rather than coronary artery involvement. Our findings suggest that the degree of laboratory abnormalities rather than the number of organ systems involved may be more useful indicators of illness severity, and may reflect differences in the degree of immune responses elicited.

Our multi-site study included children from diverse geographical regions across the United States, and captured diagnoses, laboratory values, medications and procedures. Another strength of our study is the use of LCA, which can be especially useful for identifying subgroups of individuals within a syndrome or condition who could benefit from a common intervention based on their shared characteristics. LCA is also well-suited to describing different manifestations of the novel syndrome of MIS-C. It divides patients into groups that might have been previously unrecognized, based on shared characteristics, allowing for an unbiased determination of phenotypic manifestations.^13, 17,18^ Importantly, our LCA properly accounts for between-site heterogeneity in patients by allowing different mixing proportions of subphenotypes, avoiding bias in estimated patient-level subphenotypes. Further, after identification of subphenotypes using our LCA, we also characterized the differences in variables in demographic, clinical conditions, and medications and labs, that were not used as manifest variables in our LCA. These results showed important differences in these variables across identified subphenotypes, which clinically validates our LCA results.

Our study is subject to several limitations. Given that inclusion of patients into our cohort was based on a physician’s diagnosis, we may have inadvertently included cases of suspected MIS- C who ultimately had other diagnoses, or children whose illnesses overlapped with Kawasaki disease. This risk for misclassification is low given that these diagnoses are assigned at discharge. For example, we examined Kawasaki disease diagnoses in our analyses and the prevalence was low across the three latent classes, suggesting that these patients were accurately classified. Next, our data captures hospitalizations at a PEDSnet site, and data from earlier hospitalizations at other sites was not available. More subtle symptoms from MIS-C may not have been captured in EHR data sources. Finally, the number of latent classes in LCA may be difficult to determine, which is a well-known challenge in statistical inference, and may lead to deceptive interpretations that the set of latent classes identified in an analysis represents the actual types of individuals in the population. To account for this, we have applied multiple model selection criteria in selecting the number of subphenotypes. Further, we have conducted sensitivity analyses on LCA using different numbers of subphenotypes. The final selected LCA model is a parsimonious model that describes distinct subphenotypes, which fits with clinical observations and existing literature.^19^

Our unsupervised learning methods using LCA identified a severe MIS-C subgroup with a greater extent of left ventricular dysfunction leading to shock, and need for vasopressors/inotropes. Abnormal cardiac strain has been shown to be associated with increased odds of vasoactive support requirement, duration of vasoactive support, and ICU length of stay.^20^ An association has also been shown between the degree of inflammation and cardiac injury severity, in particular CRP, troponin and NT-proBNP,^21^ as supported by our data.

Our data identified that those in the critically ill group were older and more likely to have an underlying medical condition. In the literature, children affected by MIS-C tend to have fewer underlying medical conditions, with obesity a common diagnosis if chronic conditions were present.^22^ Older age has been associated with worse outcomes from MIS-C,^23^ but the underlying immunologic mechanisms behind this observation have not yet been elucidated.

We identified a latent class subphenotype with 4-5 organ system involvement (Class 2), with overlapping features with children with acute COVID-19. A similar subphenotype has been described in earlier studies of MIS-C.^19^ These and other studies indicate the clinical overlap in some of the presentations of MIS-C and acute COVID-19. Further, the presence of abnormal strain on echocardiogram has also been associated with greater number of organ systems involved,^24^ but our findings suggest that the number of organ systems involved does not necessarily correlate with the degree of cardiac dysfunction and illness severity. Our third latent class had a milder presentation, with shorter length of stay and a lower incidence of cardiac involvement. This milder subgroup is an important cohort that warrants further exploration, as they may not require as aggressive treatment with immunomodulators to achieve the same positive outcomes.

We evaluated the proportions of each subphenotype over time, and observed a trend towards decreasing frequency of the severe MIS-C subphenotype. Earlier reports of MIS-C have indicated high levels of ICU support for patients with MIS-C. In a 2020 systematic review, Ahmed et al. reported on 39 observational studies of MIS-C patients encompassing the period January 1^st^, 2020 to July 25^th^, 2020. Out of 662 patients, 71% required admission to the intensive care unit (ICU).^25^ Further exploration of our cohort identified decreasing respiratory system involvement, cardiac involvement and shock, indicating decreased need for critical care interventions. The reasons for this decreasing illness severity over time are likely multifactorial. As clinicians’ experience with this new condition matures, earlier identification and treatment may lead to improved outcomes and decreased need for critical care. SARS-CoV-2 immunity in children has been increasing through natural infection and vaccination, rendering less children susceptible to first-time encounters with SARS-CoV-2 infection triggering MIS-C. Another hypothesis is that newer SARS-CoV-2 strains are less likely to stimulate the immune response which causes multi-system inflammation.

In conclusion, we identified a set of subgroups of MIS-C with varying illness severity, sociodemographic characteristics and organ system involvement using latent class analyses. These findings highlight the need for clinicians to recognize the varied clinical spectrum of presentations of MIS-C, and the importance of laboratory parameters to facilitate early evaluation, diagnosis and treatment.

## Data Availability

The data is not publicly available due to privacy concerns. The individual de-identified participant data will not be shared. The data that support the findings of this study may be available through request and DUA process to the corresponding authors.

## Abbreviations

LCA: latent class analyses
MIS-C: multisystem inflammatory syndrome in children
PASC: post-acute sequelae of SARS-CoV-2 infection
COVID-19: coronavirus disease 2019
SARS-CoV-2: severe acute respiratory syndrome coronavirus 2
PCR: polymerase chain reaction
EHR: electronic health record
ED: emergency department
UC: urgent care
ICU—: intensive care unit
CI: confidence intervals
ICD-10: International Classification of Diseases, version 10
PMCA: Pediatric Medical Complexity Algorithm

## Author contribution statement

Drs Chen, Jing and Liu had full access to all of the data in the study and take responsibility for the integrity of the data and the accuracy of the data analysis.

*Study concept and design:* Rao, Chen, Jing, Liu, Forrest, Maltenfort, Lorman, Schuchard

*Acquisition, analysis and interpretation of data:* Rao, Jing, Liu, Lorman, Maltenfort, Schuchard, Wu, Tong, Razzaghi, Forrest, Bailey

*Drafting of the manuscript:* Rao, Jing, Chen

*Critical revision of the manuscript for important intellectual content:* all authors

*Statistical analyses:* Jing, Liu, Chen, Wu, Tong

*Obtained funding:* Lee, Bailey, Forrest

*Administrative, technical or material support:* Rao, Chen, Bailey, Lee, Chen, Forrest

*Study supervision:* Rao, Chen, Forrest, Bailey, Forrest

