## Supplemental Material for "Clinical Subphenotypes of Multisystem Inflammatory Syndrome in Children: An EHR-based cohort study from the RECOVER program"

**Supplemental Online Content**

### **eTable 1. Definition of the variables used in latent class analysis**

| Variable | Categories | Definition |
| --- | --- | --- |
| Shock | Yes/No | - Yes, if shock^a^ is positive. - Else, no. |
| Cardiac system involvement | Coronary artery abnormalities only/myocardial dysfunction with no coronary artery abnormalities/coronary artery abnormalities and myocardial dysfunction | - Abnormality in the coronary artery only, if coronary artery dilatation or aneurysm is positive. - Abnormality not in the coronary artery, if any of the following are positive   - Heart failure (same as congestive heart failure)   - Myocarditis   - Arrhythmias   - Hypotension   - Pericarditis   - Cardiomyopathy   - Serum/plasma Troponin > 0.1 ng/mL - Both, if abnormalities in the coronary artery and not happen together. - Else, no. |
| Dermatologic system involvement | Yes/No | - Yes, if any of the following are positive   - Skin rashes   - Skin signs/symptoms   - Conjunctivitis - Else, no |
| Gastrointestinal system involvement | Yes/No | - Yes, if any of the following are positive   - Abdominal pain   - Nausea and vomiting   - Diarrhea   - Intestinal obstruction/ileus   - Gastroenteritis - Else, no |
| Hematological system involvement | Yes/No | - Yes, if any of the following are positive at any time during hospitalization   - Platelets < 150,000/microliter   - Plasma/serum D-dimer > 2 md/dL   - Lymphocytes < 1,000/microliter   - Thrombophlebitis and thromboembolism   - Aplastic anemia - Else, no |
| Neurologic system involvement | Yes/No | - Yes, if any of the following are positive   - Headache   - Cognitive signs and symptoms   - Delirium   - Encephalopathy   - Nervous system signs and symptoms - Else, no |
| Renal system involvement | Yes/No | - Yes, if any of the following are positive at any time during hospitalization   - Acute kidney injury   - Fluid and electrolyte disturbance   - Creatinine, age < 3yr, > 0.7 mg/dL OR age $\geq$ 3, > 1.0 mg/dL   - Dialysis - Else, no |
| Respiratory system involvement | Yes/No | - Yes, if any of the following are positive   - Pneumonia   - Bronchiolitis   - Bronchitis   - Cough   - Cardiorespiratory signs and symptoms   - Chest pain   - Acute respiratory distress syndrome   - Respiratory failure   - Pleurisy pleural effusion and pulmonary collapse   - Mechanical ventilation, either non-invasive or invasive - Else, no |

^a^ Codes used for shock were derived from AHRQ clusters. The codes that make up the shock cluster are found at: <https://github.com/PEDSnet/PASC/blob/main/cluster_master.csv>

**eTable 2. Proportion of patients in each class by PEDSnet site**

| Hospital | Class 1 (40.0%) | Class 2 (24.0%) | Class 3 (36.0%) |
| --- | --- | --- | --- |
| A | 33.9% | 16.3% | 49.8% |
| B | 33.1% | 34.8% | 32.1% |
| C | 54.4% | 41.4% | 4.2% |
| D | 35.5% | 23.4% | 41.1% |
| E | 37.4% | 27.0% | 35.6% |
| F | 28.0% | 15.6% | 56.3% |
| G | 40.6% | 14.6% | 44.8% |
| H | 65.4% | 21.1% | 13.5% |

### **eFigure 1. Attrition diagram of study cohort**

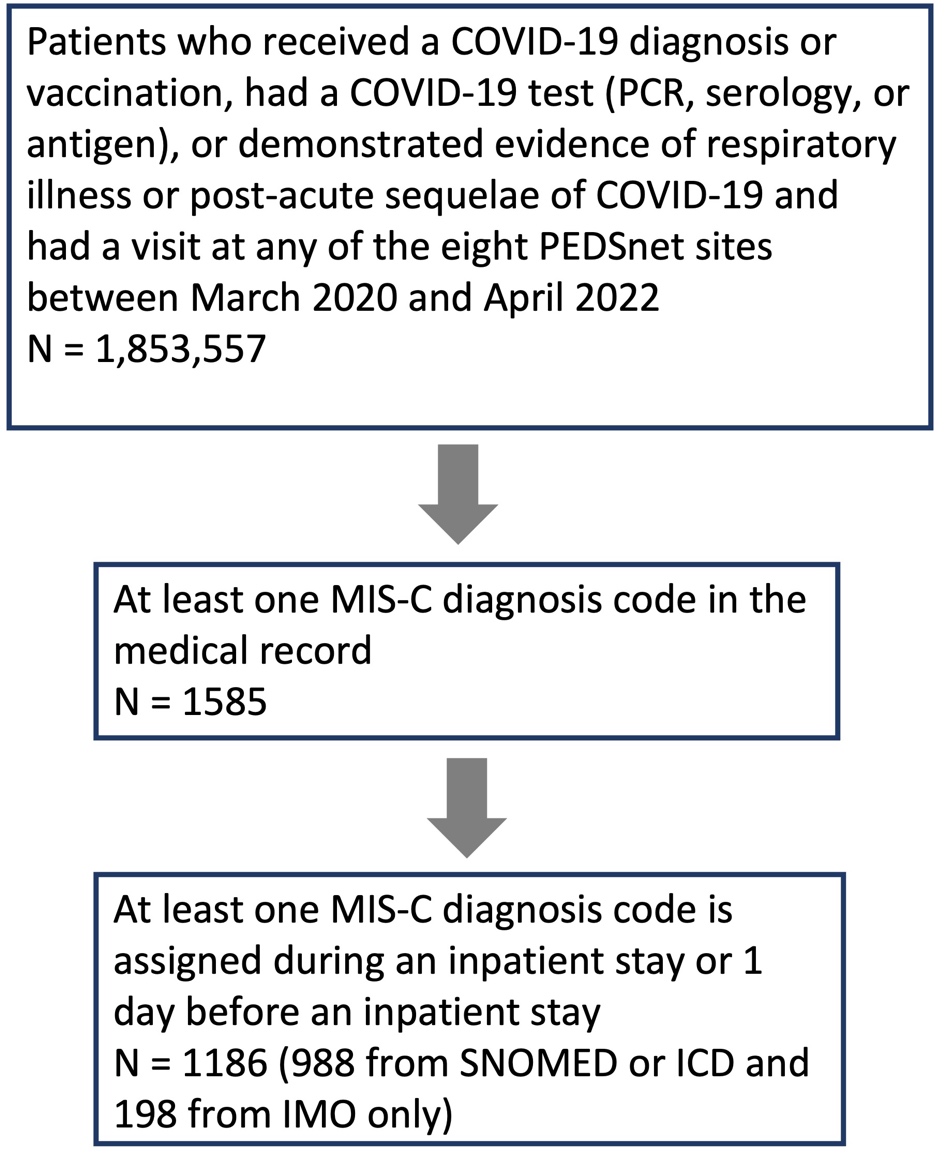

### **eFigure 2. Patient demographics and clinical characteristics by class**

**Figure legend:** Heatmap showing the prevalence of demographic variables in three classes. Each column represents a latent class, and each row represents a manifest variable. The color of the boxes represents the prevalence of variables. The legend on the top right shows the scale of the colors. Red represents prevalence close to 100% and blue represents prevalence close to 0%.

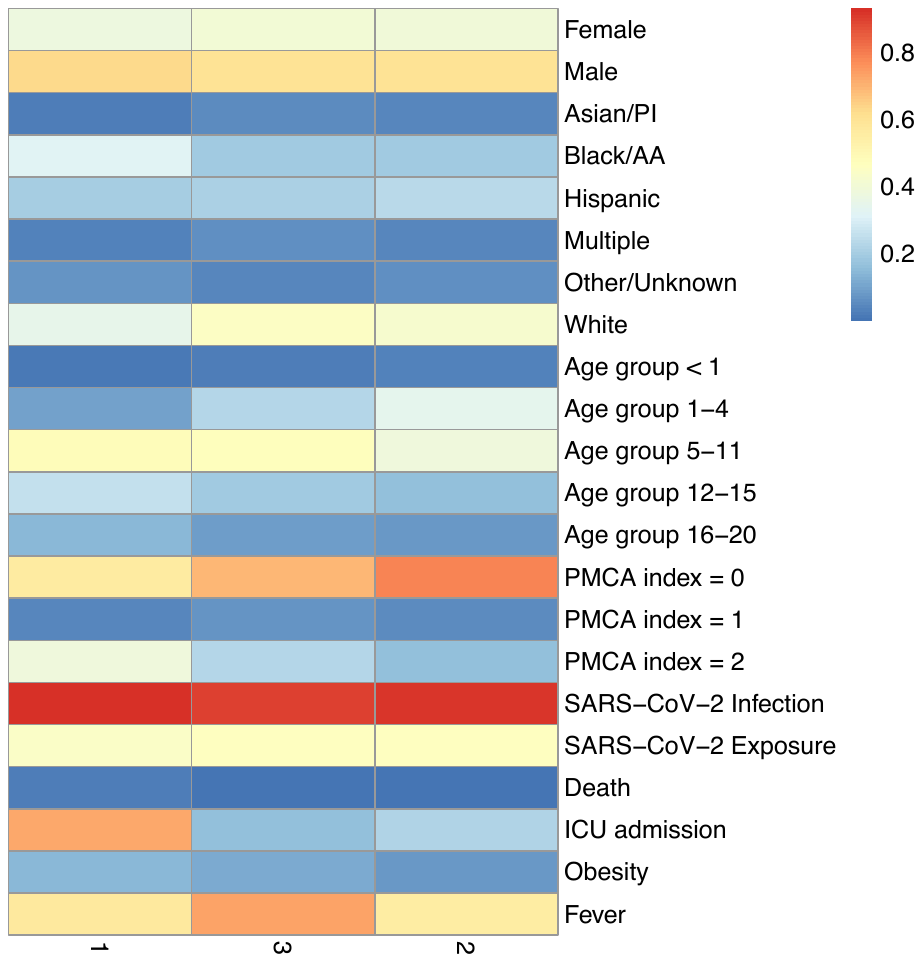

Class 1

Class 2

Class 3

**Abbreviations: PI-** Pacific islander; AA- African American; PMCA- Pediatric Medical Complexity Algorithm; SARS-CoV-2 - severe acute respiratory syndrome coronavirus 2; ICU- intensive care unit

### **eFigure 3. Patient characteristics of conditions, laboratory results, and medications in three latent classes**

**Figure legend**: The I-squared statistic measures the heterogeneity among the latent classes. P-values are obtained through Cochran’s Q test, where the null hypothesis is no between-class heterogeneity. A large I-squared and a small p-squared indicate a large between-class heterogeneity. The first panel on the right shows the estimated prevalence and its 95% confidence intervals. **A)** Variables related to cardiac, skin, gastrointestinal, and hematological systems. **B)** Variables related to neurological, renal, respiratory systems, and Kawasaki disease. **C)** Medications.

**Skin**

**Gastrointestinal**

**Hematological**

**Cardiac**

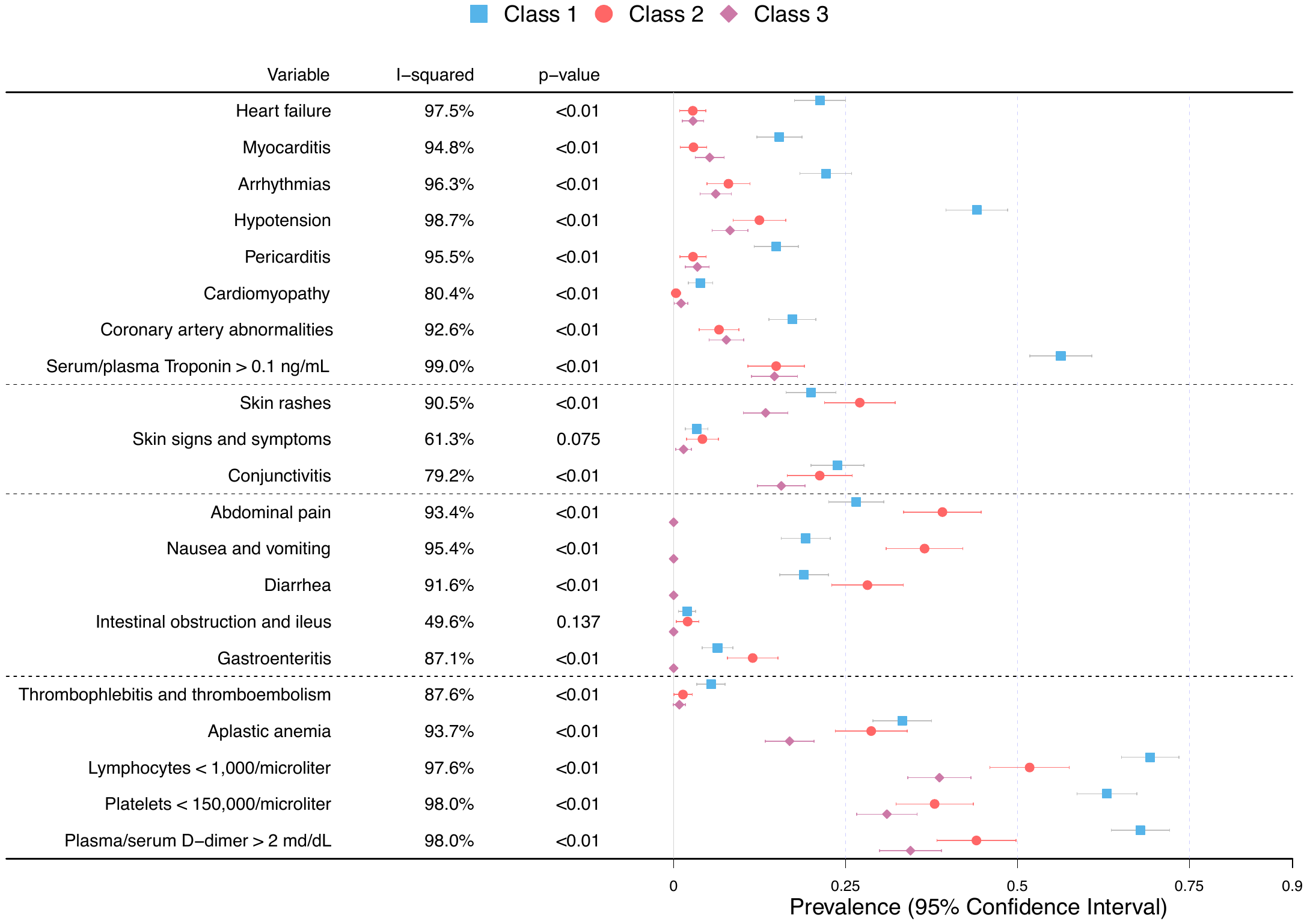

A

**Medications**

C

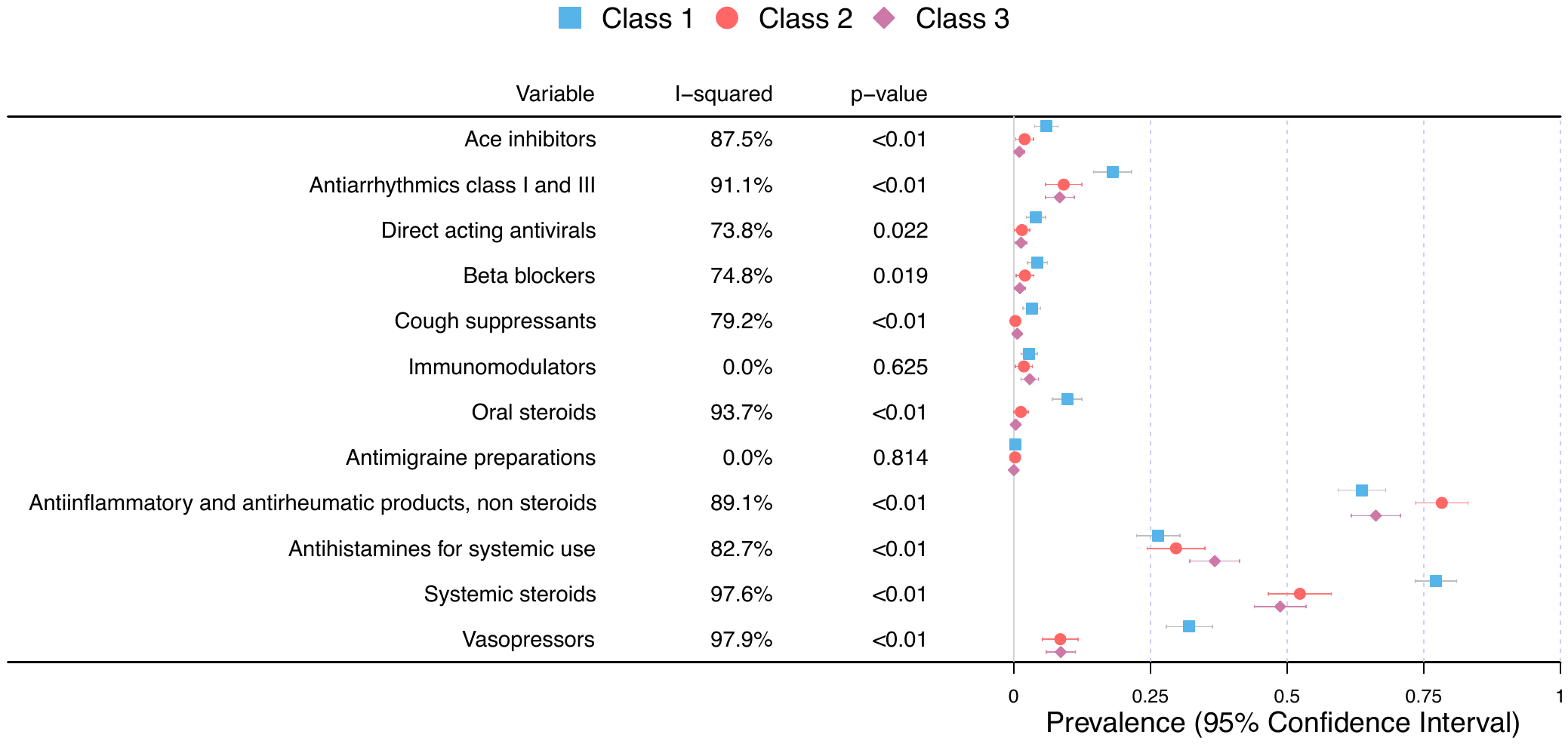

**Renal**

**Respiratory**

**Other**

**Neurological**

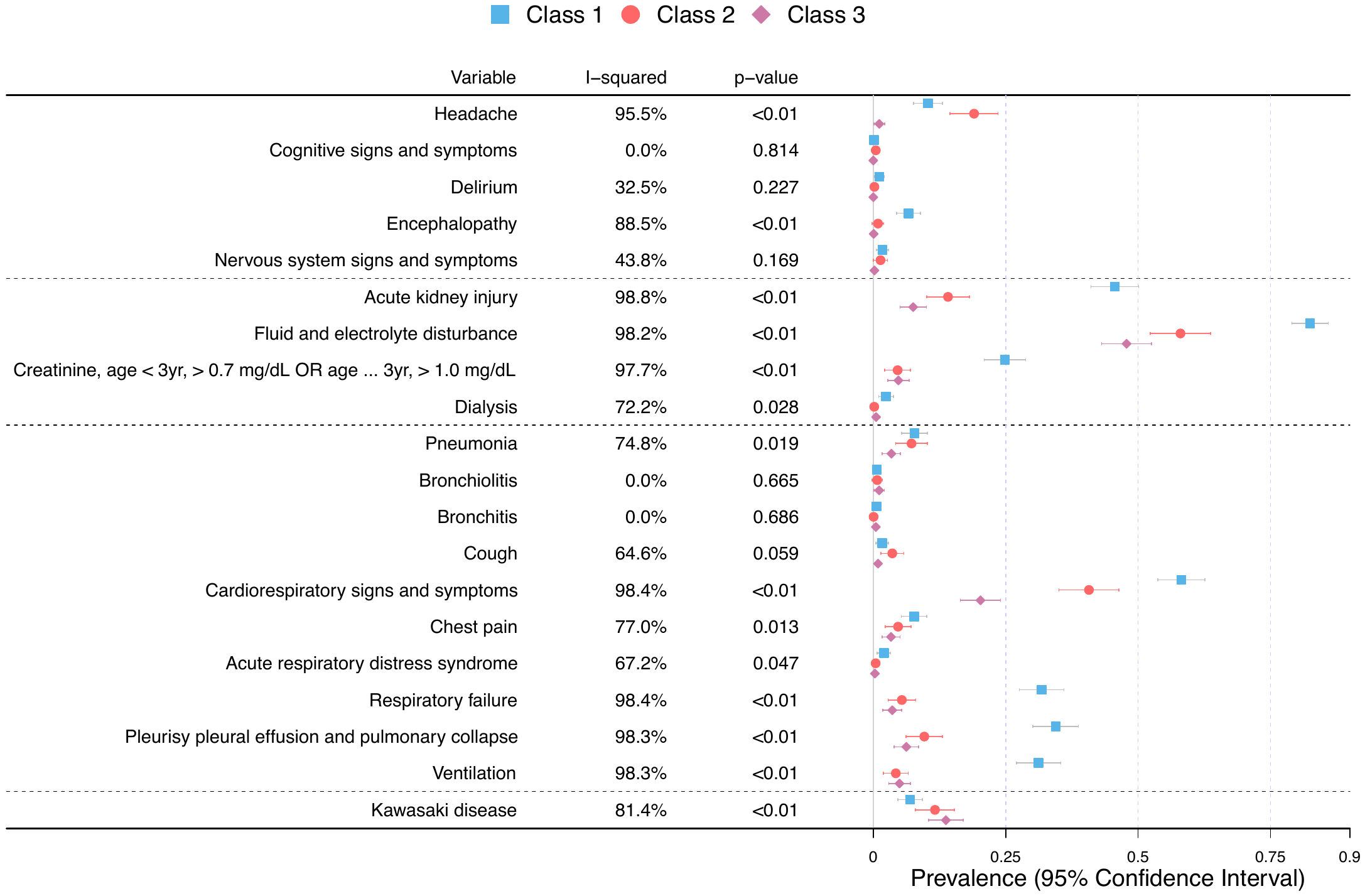

B

### **eFigure 4. Comparison of latent classes in children testing positive for SARS-CoV-2 with children with MIS-C**

**Figure legend:** Heatmaps showing the latent classes and characteristics of children from two subpopulations **A)** children testing positive for SARS-CoV-2 and **B)** children with MIS-C. Each column represents a latent class, and each row represents a manifest variable. The color of the boxes represents the prevalence of variables. The legend on the top right shows the scale of the colors. Red represents prevalence close to 100% and blue represents prevalence close to 0%. **A) Latent classes of COVID-19.** We applied LCA with three latent classes to 59,740 patients testing positive for SARS CoV-2 by PCR who were not diagnosed with MIS-C. The variables used were the same as in MIS-C LCA. The evaluation time of the variables is -7-28 days from the first positive testing date. The proportions of Class 1, 2, and 3 were 1.7%, 12.8%, and 85.6%. **B) Latent classes of MIS-C** as shown in **Figure 2**.

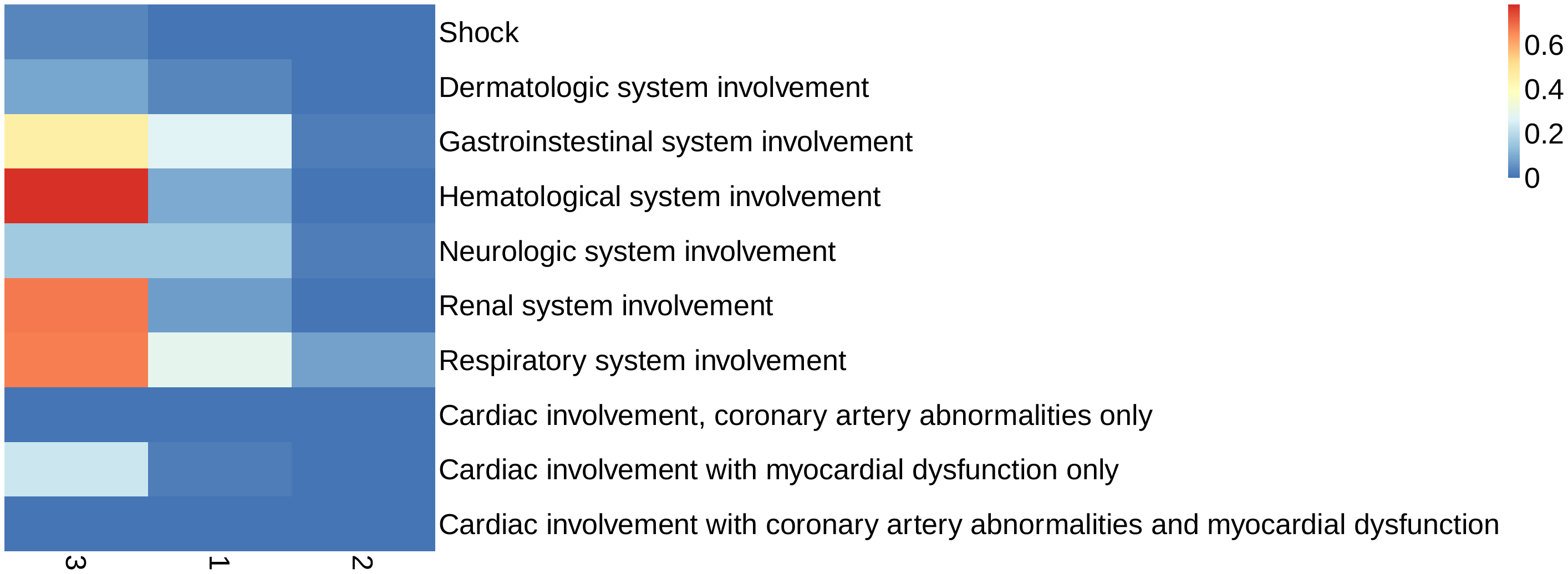

Class 1

Class 3

Class 2

A

B

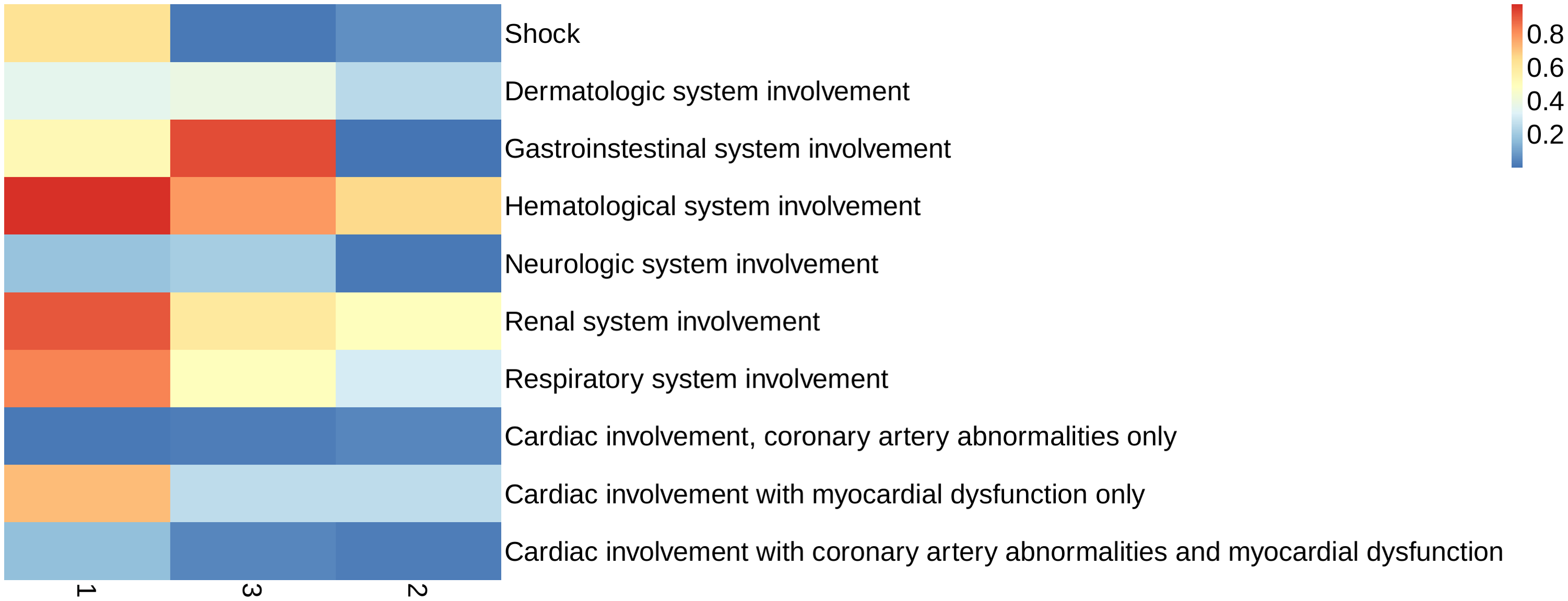

Class 1

Class 2

Class 3

### **eFigure 5. 2-Dimensional plot comparing the distance between MIS-C and COVID-19 PCR positive subpopulations**

**Figure legend:** Patients were grouped into latent classes (subpopulations) according to their maximum posterior class membership probabilities. Fixation index (Fst), a measure of population distance, was calculated for each pair of subpopulations using the eight variables in LCA. To visualize the distances between subpopulations, we applied the multidimensional scaling technique to map the distances onto a 2-dimensional plot. Closer subpopulations have larger similarities. For example, MIS-C Class 2 is similar to COVID-19 Class 1, while MIS-C Class 1 is distinct from other subpopulations.

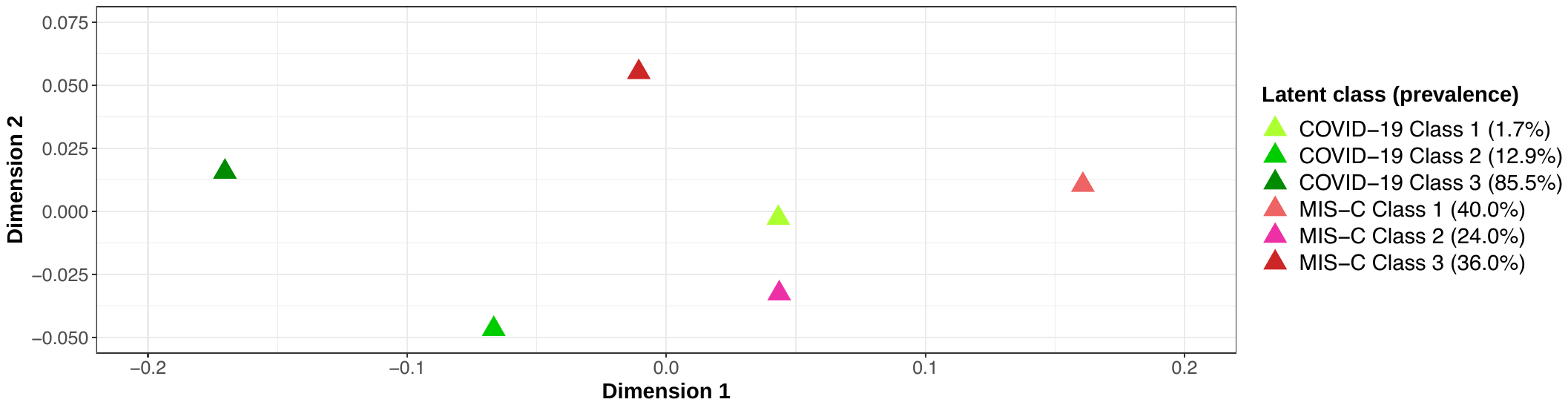

### **eFigure 6. Prevalence of organ system involvement among children with MIS-C over time, from March 2020 to April 2022**

**Footnote:** The calendar time is segmented quarterly. Time intervals with small sample sizes are combined and labeled by a star. Statistically significant changes over time were calculated using the Chi Square test.

^a^ p values < 0.05

^b^ p values < 0.01

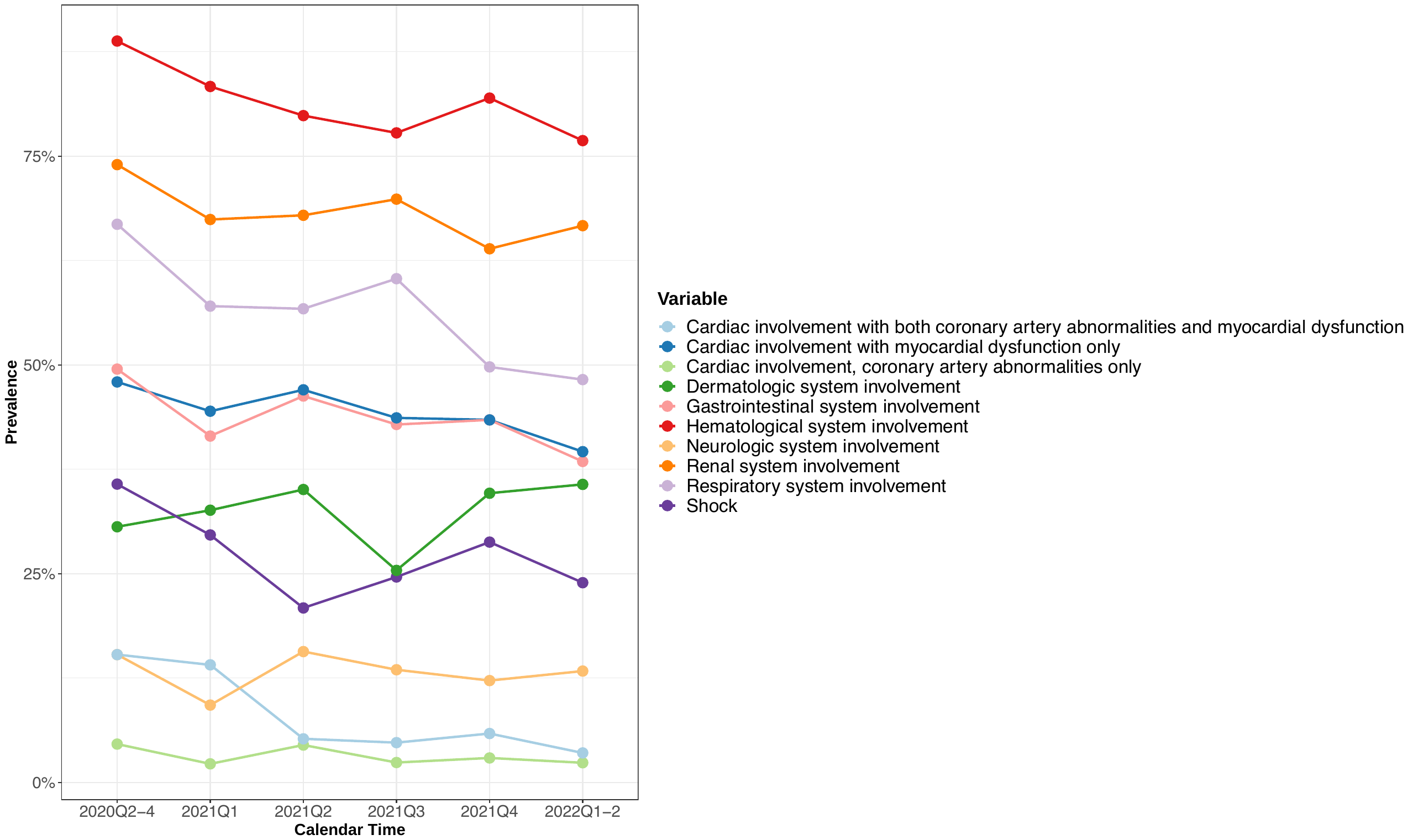

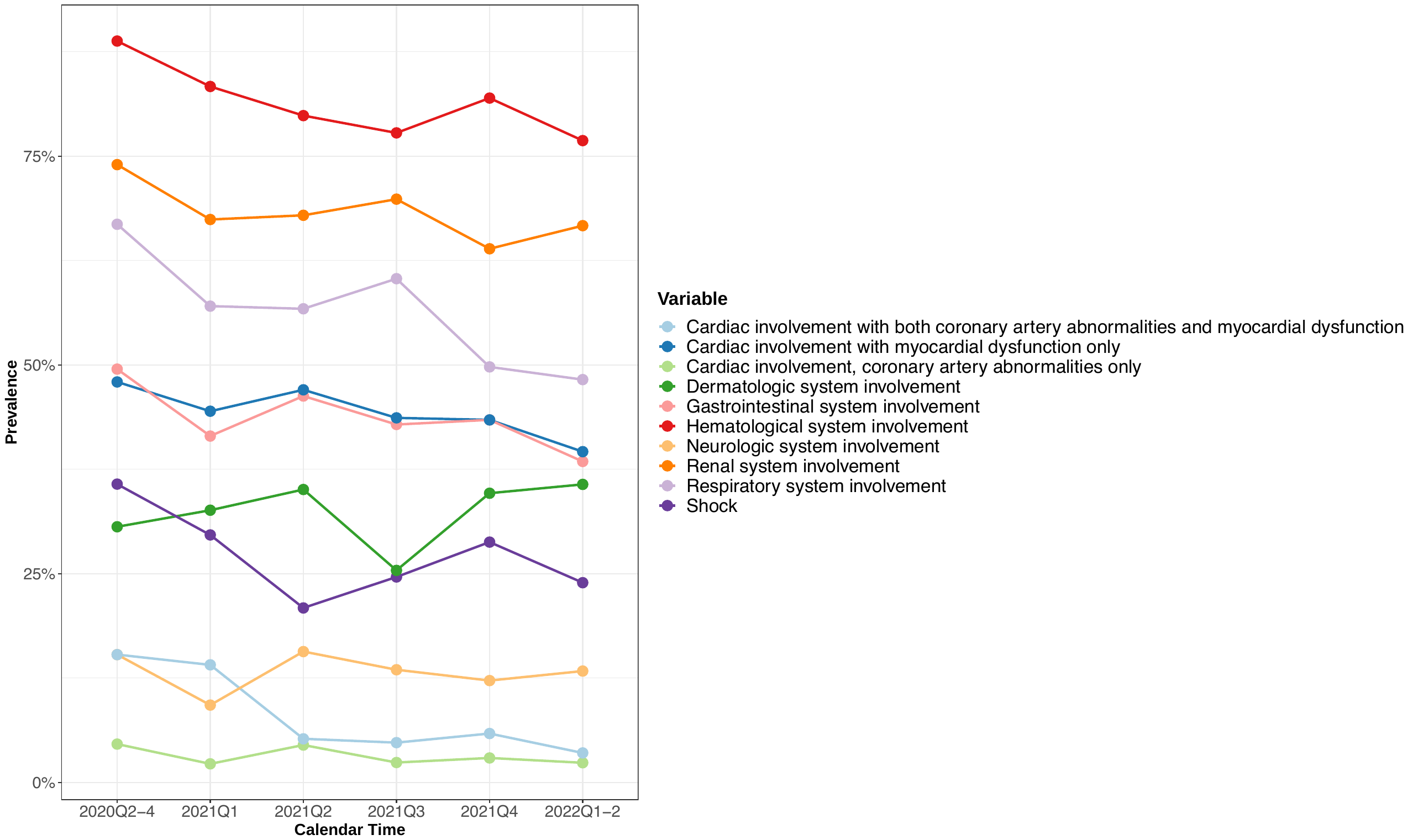

*

*

a

a

b

b

### **eFigure 7. Latent Class Analyses using 4 classes**

**Figure legend: A)** Heatmap showing the characteristics of the four latent classes. Each column represents a latent class, and each row represents a manifest variable. The color of the boxes represents the prevalence of variables. The legend on the top right shows the scale of the colors. Red represents prevalence close to 100% and blue represents prevalence close to 0%. **B)** The proportion of patients in each class by site.

A

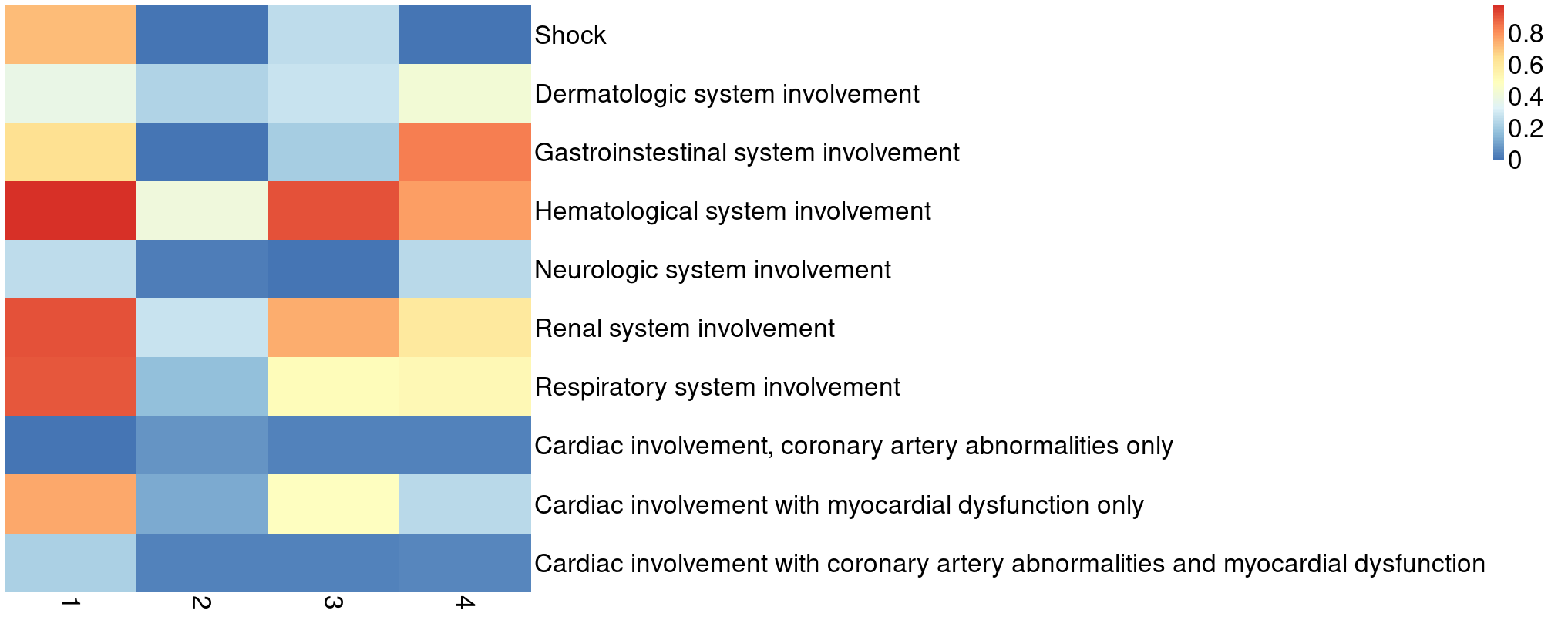

Class 4

Class 3

Class 2

Class 1

| Site | Class 1 25.8% | Class 2 16.3% | Class 3 34.7% | Class 4 23.1% |
| --- | --- | --- | --- | --- |
| A | 15.9% | 23.5% | 43.8% | 16.7% |
| B | 21.9% | 11.9% | 32.6% | 33.6% |
| C | 50.5% | 1.9% | 3.7% | 43.9% |
| D | 23.7% | 20.6% | 32.8% | 23.0% |
| E | 26.4% | 18.8% | 29.1% | 25.7% |
| F | 23.1% | 50.2% | 12.2% | 14.6% |
| G | 17.4% | 12.5% | 58.3% | 11.8% |
| H | 53.2% | 5.4% | 16.8% | 24.6% |

B

### **eFigure 8. Latent Class Analyses using 5 classes**

**Figure legend: A)** Heatmap showing the characteristics of the five latent classes. Each column represents a latent class, and each row represents a manifest variable. The color of the boxes represents the prevalence of variables. The legend on the top right shows the scale of the colors. Red represents prevalence close to 100% and blue represents prevalence close to 0%. **B)** The proportion of patients in each class by site.

A

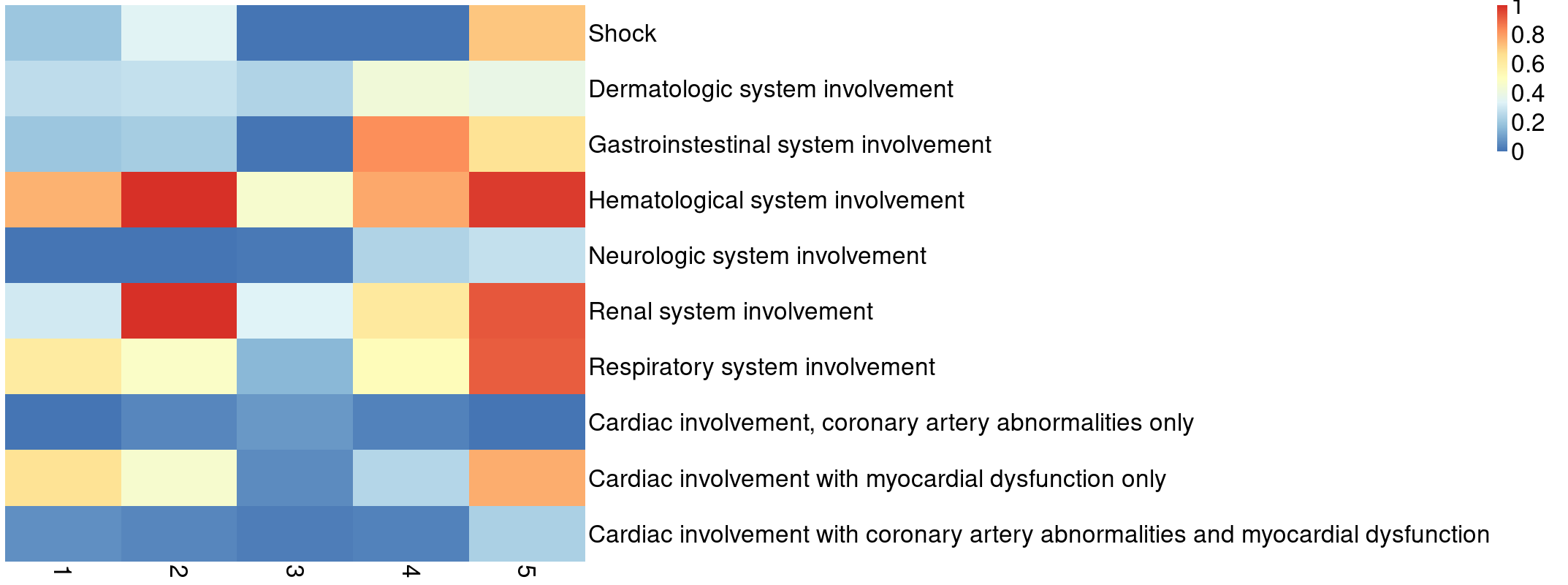

Class 5

Class 4

Class 3

Class 2

Class 1

| Site | Class 1 18.5% | Class 2 21.5% | Class 3 24.4% | Class 4 25.4% | Class 5 10.2% |
| --- | --- | --- | --- | --- | --- |
| A | 11.6% | 32.5% | 24.4% | 17.3% | 14.2% |
| B | 6.2% | 22.2% | 14.6% | 35.6% | 21.4% |
| C | 3.2% | 0.8% | 2.0% | 44.2% | 49.9% |
| D | 3.3% | 24.3% | 24.4% | 24.4% | 23.6% |
| D | 3.3% | 24.3% | 24.4% | 24.4% | 23.6% |
| E | 32.0% | 2.3% | 13.9% | 24.7% | 27.0% |
| F | 12.7% | 8.2% | 45.2% | 12.8% | 21.0% |
| G | 8.8% | 43.8% | 17.9% | 13.6% | 15.9% |
| H | 34.6% | 0.0% | 0.0% | 17.4% | 48.0% |

B

### **eTable 3. Model selection criteria of Latent Class Analysis using 3, 4, and 5 classes.**

**Figure legend:** The values of log-likelihood and Bayesian Information Criterion (BIC) with 2-6 latent classes. The best BIC is in red and labeled by a star (A lower BIC indicates a better model).

| Number of latent classes | 2 | 3 | 4 | 5 | 6 |
| --- | --- | --- | --- | --- | --- |
| Log-likelihood | 5754.58 | 5703.67 | 5667.42 | 5651.43 | 5638.64 |
| BIC | 11657.81 | **11633.85*** | 11639.21 | 11685.10 | 11737.36 |
